# Understanding the present status and forecasting of COVID―19 in Wuhan

**DOI:** 10.1101/2020.02.13.20022251

**Authors:** Toshihisa Tomie

## Abstract

The present status of COVID―19 is analyzed and the end of the disease is forecasted. The peak of the epidemic is different in three regions, Wuhan, Hubei province except Wuhan, and mainland China except Hubei. In two regions except Wuhan, the peak of the epidemic passed ten days ago. If the trend until February 11 does not change, the disease may end by the end of February. In Wuhan, the epidemic reached a peak but the reported number of newly infected patients fluctuates largely. We need to know the reason for the big fluctuation to forecast the end of the disease.

## 1. Introduction

Pneumonia originated in Wuhan, which is referred as COVID―19 in the following, is now a great fear in the world. The fear of people is caused by no existence of forecasting of the final scale of the disaster and the end of the disease. This paper reports forecasting of the end of the COVID―19 epidemic. As a result of the data analysis, it was found that the peak of the number of newly infected people differed by region, and the peak was February 2 in China outside Hubei Province. The decay constant was about 8 days. On February 11, the number of newly infected people reduced to less than half of the peak value. In Wuhan city, the source of the virus, the peak was reached, but the fluctuation of the number of new infections is large. The convergence of fluctuation is necessary to forecast the end of the epidemic in Wuhan.

### 2. Reference of infectious disease outbreak

In order to understand the behavior of the general epidemic, the influenza epidemic in Japan over the past decade is shown in Fig. 1 (ref. 1). The duration of influenza epidemics was four to six weeks. To avoid complexity, epidemics of three years are shown in Fig. 2. An epidemic can be described by a Gaussian around the peak of the epidemic. The 2019 epidemic, which is referred as JpnInf2019 in the following, has the smallest deviation from a Gaussian and has a shorter epidemic period. As shown by a solid line in Fig. 2, the epidemic of JpnInf2019 can be described by a Gaussian with a peak at 3.65 week and the number of new patients of 56 people at fixed observatories of diseases, and the “decay constant” which is the time duration between the peak and the time when the number of new infections decreases to 1/e of the peak value is 2.6 weeks. Although the full width at half maximum (FWHM) is common as a descriptive term for a Gaussian, the term “decay constant” is used to simplify the formula. FWHM equals to 1.67 × “decay constant”.

**FIG.1:**
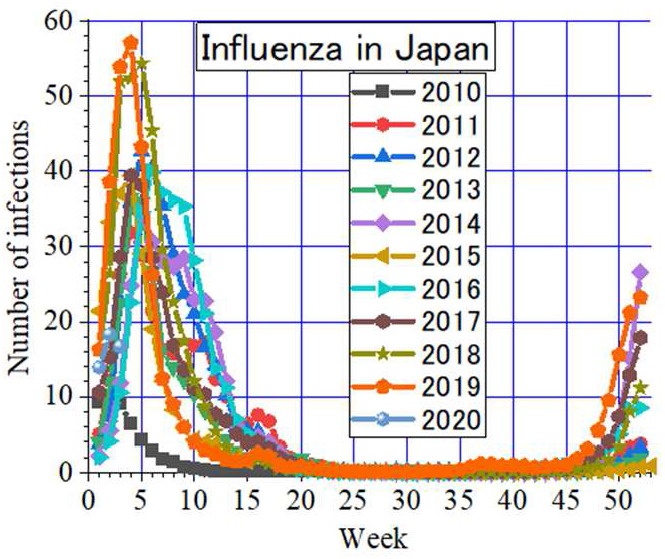
Epidemic of influenza in Japan over the past 10 years.

**FIG.2:**
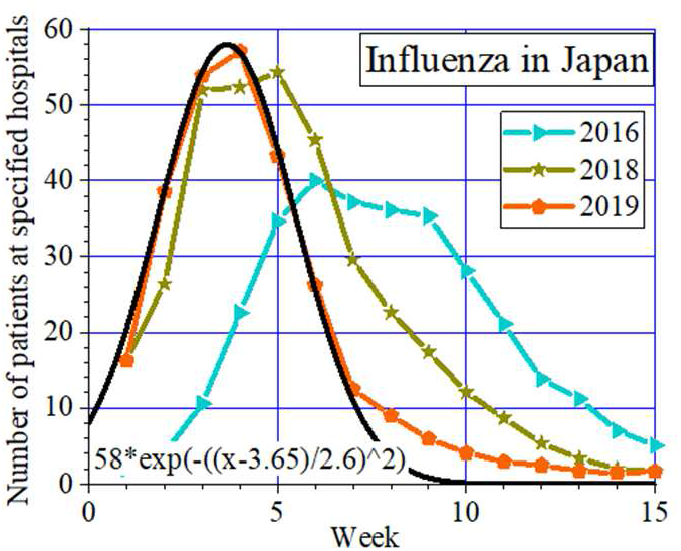
Epidemic of the infectious disease can be fitted with a Gaussian.

### 3. Analysis of the outbreak of COVID―19

The data on the epidemic of COVID―19 was obtained from ref. 2.

The horizontal axis is the number of days since the beginning of the year; January 1 is 1 and February 1 is 32. There are three fitting parameters: peak position, peak value, and decay constant. By assuming the decay constant to be 18 days which was that of JpnInf2019, the reported data points are fitted well by a Gaussian with a peak of February 27 and a peak value of 20,000 people, as shown in Fig. 3. The cumulative number of infected people is well reproduced by the curve, as shown in Fig. 4.

**FIG.3:**
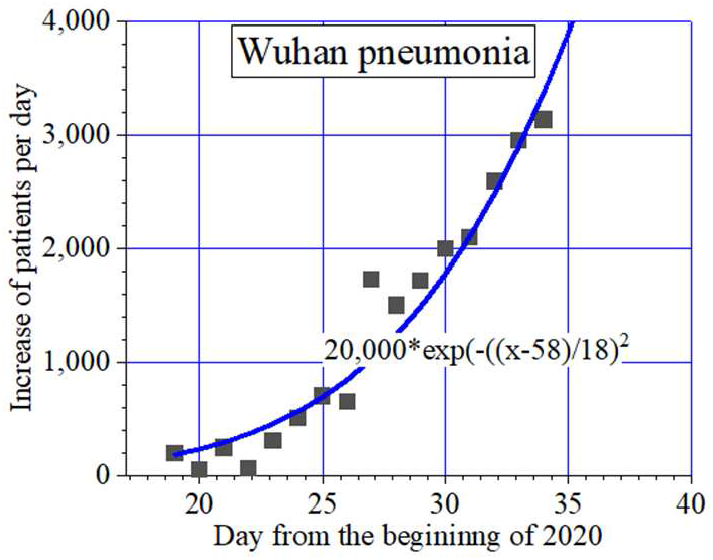
Fitting of COVID―19 with a Gaussian by fixing the decay constant to be 18 days.

**FIG.4:**
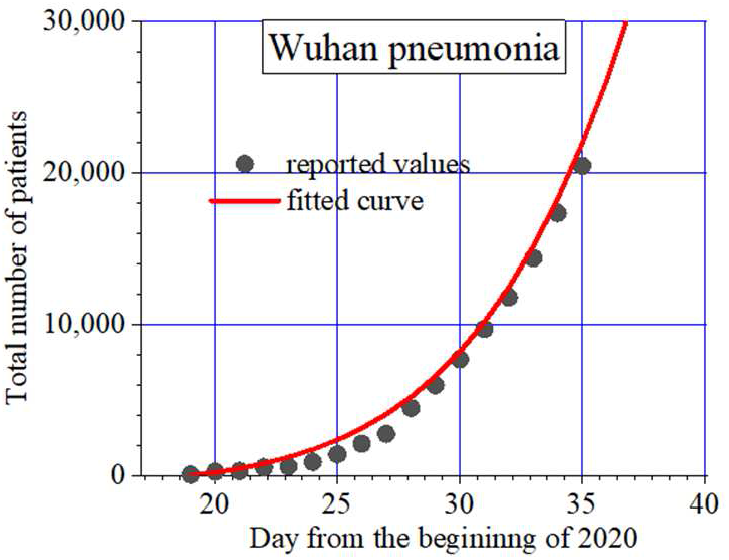
Comparison of reported values with the fitted Gaussian.

The number of newly infected people expected by the fitting curve up to 100 days is shown in Fig. 5 and the expected cumulative number of infected people is shown in Fig. 6. If the epidemic follows the fitting curve, the total number of infected people will eventually exceed 600,000 people.

**FIG.5:**
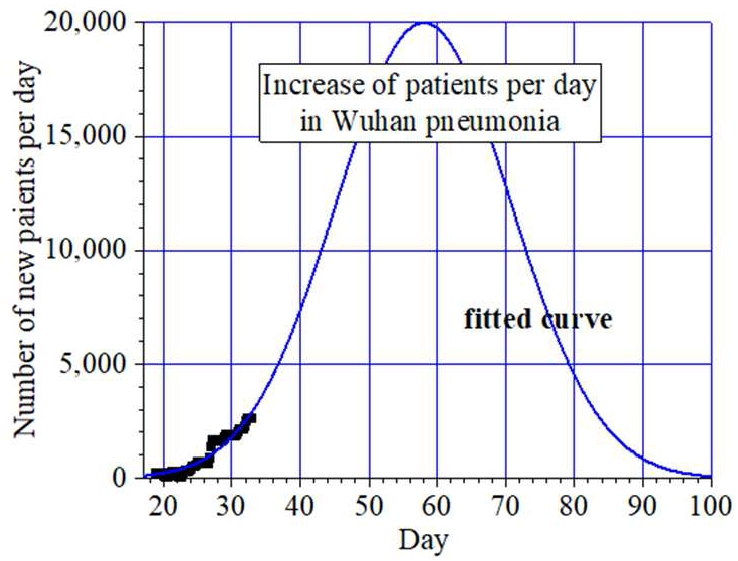
Increase of patients per day when epidemic of COVID―19follows the curve.

**FIG.6:**
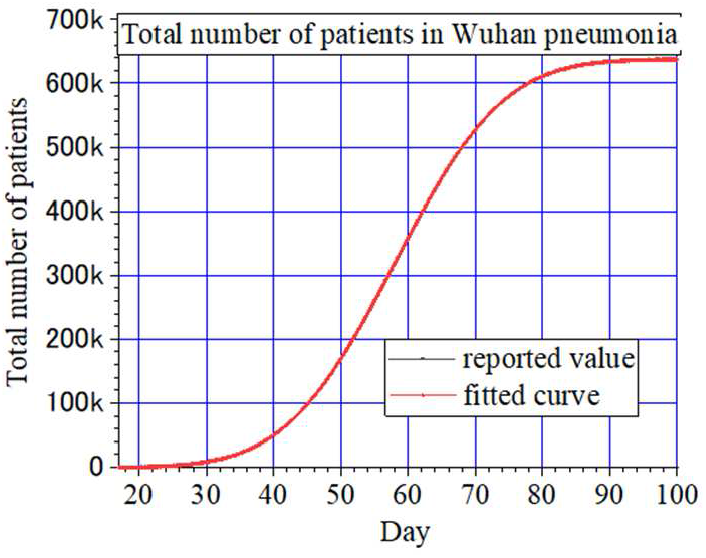
Total number of patients may reach 60,000 if the disease follows the curve.

### 4. Deviation of the data from the fitted Gaussian from February 6^th^

The above fitting was carried out on February 2. For three days from February 3 to 5, the number of newly infected people followed the predicted curve as shown by red circles in Fig. 7. However, the deviation of the data from the fitting curve from February 6 increased as shown by blue circles. Fitting these data with a simple Gaussian became difficult. The deviation of an epidemic of disease from a Gaussian is a mystery. This mystery was solved by a new analysis described below.

**FIG.7:**
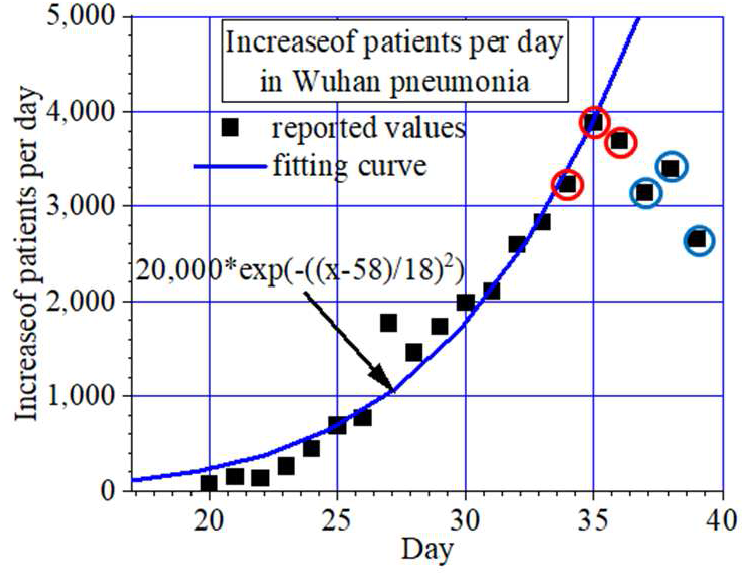
Number of newly infected patients deviated from the fitting curve from Feb. 6.

### 5. Analysis divided into three regions

In the above, we dealt with the data from all over mainland China to minimize the statistical fluctuation. However, social conditions differ greatly depending on the region. For example, the city of Wuhan has been blocked from outside since January 23. In addition, although the rush construction of new hospitals has been carried out, owing to an extremely large number of potential infections, there is a possibility that the ability of the examination and treatment by medical institutions has not caught up. These circumstances cause a serious deviation of the reported number of infected people from the actual and can also affect the epidemic. In order to analyze in more detail, the data of the pneumonia was also obtained from the website of the Hubei government (ref.3), which publishes data of the entire Hubei province and Wuhan city. Using these newly acquired data, the epidemic was separately analyzed in three regions: Wuhan city, Hubei Province except Wuhan, and whole China except Hubei Province. The results are shown in Fig. 8.

**FIG.8:**
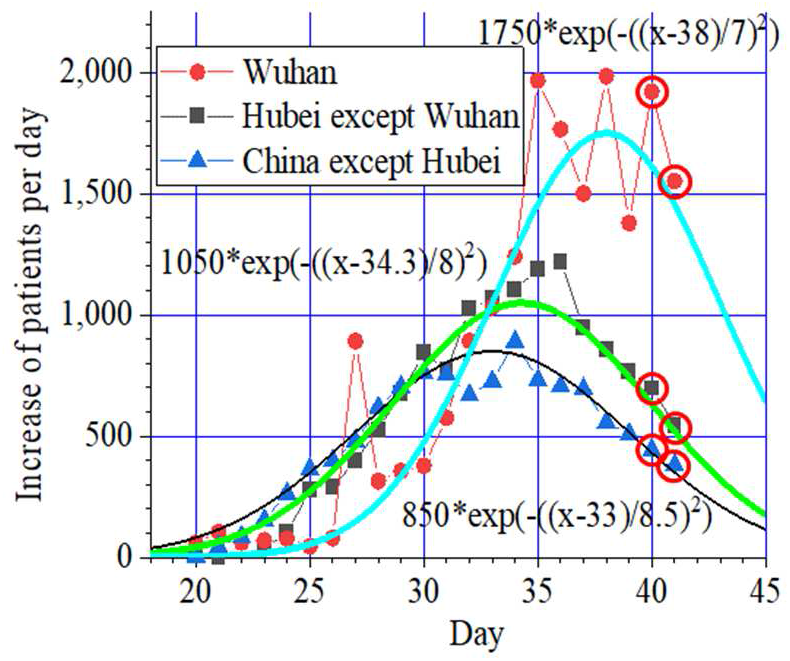
In three regions, the peak of the number of newly infected patients are different. The decay constant of the fitting Gaussians is 8 days.

By dividing China into three regions, the mystery of the non-Gaussian change of the data points seen in Fig. 7 was solved. The days of the peaks in the three regions were different. In the whole China except Hubei Province, the peak was on February 2, the peak for the Hubei province except Wuhan was February 3, and Wuhan has a peak around February 7. Because the three Gaussians of similar sizes with different peak positions were summed-up, it was not possible to fit the data by a single Gaussian. Our success gives an important hint for future analysis of other infectious diseases. Separating the data into several groups may clarify factors affecting an epidemic.

The decay constant revealed in the new analysis was about 8 days, which is less than half of that of JpnInf2019. Fitting shown in Fig. 8 was performed using the data up to February 9. The data on the subsequent two days followed the curves as shown by red circles. The number of new patients on February 11 was smaller than half of the values at the peaks in two regions except Wuhan city.

In the case of JpnInf2019 shown in Fig. 2, at the skirt part, the epidemic behavior changed from a Gaussian to exponential decay. Therefore, except in Wuhan city, around February 20 the number of newly infected people will be further reduced to 1/e. If the present trend does not change, the number of new infections is likely to be negligibly small by the end of February.

In Wuhan, the source of the virus, the peak was reached but the daily fluctuation is very large. It is necessary to understand the cause of the big fluctuation in forecasting the end of the epidemic in Wuhan.

## 6. Summary

The epidemic of COVID―19 was analyzed. When the fitting was carried out on February 2 by assuming the decay constant of 18 days as was observed in JpnInf2019, a relatively good fitting curve was obtained. However, after February 6^th^, fitting by a single Gaussian became difficult. Therefore, we divided the data into three regions; Wuhan, Hubei Province except Wuhan, and mainland China other than Hubei Province, and it was found that the peak days in the three regions were different. The decay constant revealed in the new analysis was about 8 days. Outside Wuhan, the number of new infections has already decreased to less than half of their peak value on February 11. It is expected that the number of new infections will be negligibly small by the end of February if the present trend does not change. In Wuhan, although the number of new infections reached its peak, the fluctuation of the new infections is large. It is necessary to understand the cause of the big fluctuation for forcasting the end of the epidemic.

## Data Availability

All the data used in the manuscript are available from references.

## Notes

### Competing Interest Statement

The authors have declared no competing interest.

### Funding Statement

No external funding was received

